# Immune Markers Are Associated with Cognitive Performance in a Multiethnic Cohort: the Northern Manhattan Study

**DOI:** 10.1101/2021.01.18.21250068

**Authors:** Mitchell S. V. Elkind, Michelle Moon, Tatjana Rundek, Clinton B. Wright, Ken Cheung, Ralph L. Sacco, Mady Hornig

## Abstract

**OBJECTIVE:** To determine whether immune protein panels add significant information to correlates of cognition.

**BACKGROUND:** Immune mechanisms in vascular cognitive impairment and dementia are incompletely characterized.

**DESIGN/METHODS:** A subsample of the prospective Northern Manhattan Study underwent detailed neuropsychological testing. Cognitive scores were converted into Z-scores and categorized into four domains (memory, language, processing speed, and executive function) based on factor analysis. Blood samples were analyzed using a 60-plex immunoassay. We used least absolute shrinkage and selection operator (LASSO) procedures to select markers and their interactions independently associated with cognitive scores. Linear regression models assessed cross-sectional associations of known correlates of cognition with cognitive scores, and assessed model fit before and after addition of LASSO-selected immune markers.

**RESULTS:** Among 1179 participants (mean age 70±8.9 years, 60% women, 68% Hispanic), inclusion of LASSO-selected immune markers improved model fit above age, education, and other risk factors (p for likelihood ratio test<0.005 for all domains). C-C Motif Chemokine Ligand 11 (CCL 11, eotaxin), C-X-C Motif Chemokine Ligand 9 (CXCL9), hepatocyte growth factor (HGF), and serpin E1 (plasminogen activator inhibitor-1) were associated with each of the domains and with overall cognitive function. Immune marker effects were comparable to conventional risk factors: for executive function, each standard deviation (SD) increase in CCL11 was associated with an effect equivalent to aging three years; for memory, HGF had twice the effect of aging.

**CONCLUSIONS:** Immune markers associate with cognitive function in a multi-ethnic cohort. Further work is needed to validate these findings and determine optimal treatment targets.

## Introduction

Communication between the brain and immune system is increasingly recognized as critical for brain health and neuroprotection after cerebrovascular, metabolic and microbial challenges.^1,2,3,4^ Questions remain, however, regarding the role of inflammation in vascular and neurodegenerative mechanisms of cognitive decline, mild cognitive impairment (MCI) and dementia; the specific inflammatory pathways that are most important;^5,6^ and the role of inflammation in populations, particularly Hispanics, with a high burden of vascular risk factors but that have been underrepresented in research.^7,8,9^ Innate immunity, activation of complement and microglia, and humoral and cellular immunity may all contribute.^10,11,12,13^ Much of this work, moreover, is based on preclinical models or small sample sizes in humans.

We hypothesized that a multiplex immunoassay measuring 60 immune molecules would provide insight into pathways involved in cognitive decline. We explored this hypothesis in a population-based cohort study, the Northern Manhattan Study (NOMAS), a sample of Hispanic and non-Hispanic participants from New York City.

## METHODS

### Study Population

NOMAS is a racially/ethnically diverse prospective cohort study consisting of 3,298 stroke-free participants enrolled between 1993 and 2001. The study was initially designed to evaluate effects of medical, socio-economic, and vascular risk factors on incidence of stroke and other vascular outcomes. Eligible participants were individuals residing in northern Manhattan for ≥ 3 months in a household with a telephone, with no prior history of stroke, and age ≥40 years old at the time of enrollment. Subjects were identified by random-digit dialing, and in-person interview and assessment were conducted by trained bilingual research assistants. A subcohort of 1290 participants included 1091 subjects remaining clinically stroke-free from the original cohort as well as a sample of unrelated household members (n=199). MRI and detailed neuropsychological assessments were conducted, and blood samples were collected; methods of recruitment and characteristics of this sample have been described previously.^14^ The study was approved by the Columbia University and University of Miami Institutional Review Boards, and all participants provided informed consent.

### Data collection

Demographics and risk factors were collected through in-person interviews, as described previously.^14^ Race-ethnicity was determined by self-identification. Insurance status was defined no insurance or Medicaid versus private insurance / Medicare^23^. Educational achievement was self-reported as number of years in school and degree achieved. Physical activity was evaluated by in-person questionnaire^35^ and defined dichotomously. Smoking was categorized as current (within the past year), former, or never smoker of cigarettes, cigars or pipes. Standardized questions adapted from the Behavioral Risk Factor Surveillance System assessed hypertension and diabetes. Hypertension was defined either as participant self-report of hypertension, blood pressure measurement of 140/90 mmHg or greater, or use of anti-hypertensive medication. Diabetes was defined either as participant self-report of diabetes, fasting glucose of 126 mg/dL or greater, or use of insulin or oral anti-diabetic medications.

### Cognitive assessment

Neuropsychological assessments were administered by trained bilingual research staff in English or Spanish, as previously described.^15^ We developed cognitive domain-specific scores to enable interpretability of cognitive functions (Table e-1). Because of lack of population norms for older cohorts, we used baseline neuropsychological scores as our cohort norms.^16^ To achieve comparability, neuropsychological tests were standardized against cohort-specific normative values. For timed tests (Color Trail 1, Color Trail 2 and Grooved Pegboard), because some participants did not complete tasks within the maximum time allowed, we calculated their new “imputed” time by adding penalty time to the maximum time allowed. The penalty time was obtained by multiplying the number of incomplete items by “expected time per correct items completed” estimated using Kaplan-Meier methods. The standardized scores for individual tests were then categorized into four key cognitive domains (memory, language, processing speed and executive function) based on an exploratory factor analysis and prior findings.^16^ Thereafter, cognitive domain-specific scores were calculated by taking the mean of the individual standardized test scores selected for each domain. Cognitive outcomes were domain-specific z scores (memory, language, processing speed and executive function) at baseline and a composite cognitive function score obtained by averaging those four domain scores.

### Multiplex immunoassay assessment

We completed assays of 60 immune molecules in 1179 participants from stored fasting plasma drawn at the time of a detailed cognitive assessment.^17,18^ We used a customized, highly multiplexed, magnetic bead-based immunoassay.^19,20^ Our assay included molecules integral to inflammatory responses, reflecting relative activation of Th1 (pro-inflammatory), Th2 (counter-regulatory, autoimmunity-promoting), Th17 (innate mucosal anti-pathogen defenses, including in gut), T regulatory, B (antibody-producing) and NK cells, and, more generally, innate and adaptive (memory) arms of the immune system; chemoattractants (chemokines); growth factors (including those vital to vascular remodeling); neurotrophic factors; cell adhesion markers; and adipokines.

Plasma samples were coded, randomized and assayed in duplicate. Median fluorescence intensities (MFI) of each analyte-specific immunoassay bead set were detected by the flow- and fluorescence-based Luminex 200™ detection platform (Luminex Corporation, Austin, TX).^21^ Data were processed using a quality control (QC) algorithm that calibrates performance of an expanded set of serial standard curves and an in-house plasma control run on every assay plate, monitoring intra- and inter-plate covariance (CV) and bead counts. Only plates with mean intra-assay %CV <15% were accepted. Samples failing to meet QC criteria were designated for re-run when feasible (%CVs >25%, bead counts <30). MFI values exceeding machine limits of reliable detection (>25,000) were excluded. Because interpolated concentrations can introduce bias for samples with very low or high values in relation to the serial standard curves, averaged MFI values for all 60 cytokines were used in the final statistical analyses rather than absolute concentrations.^17,18^ The final data set (test samples) had mean intra-assay %CV of 4.6% (standard deviation [SD] 2.2%) and 0.5% of all possible intra-assay %CV values were >25%, across all cytokines. Per-cytokine intra-assay %CV values ranged from 1.9% to 7.9%.

### Statistical analyses

Distributions of socio-demographic characteristics and vascular risk factors for the study cohort were calculated.

Association between cognitive outcomes and immune markers was assessed using a two-step hybrid Least Absolute Shrinkage and Selection Operator (LASSO) approach^22^: first, we performed the LASSO technique to select a subset among the 60 immune markers (logarithm base 2-transformed and standardized) and all possible two-way interactions that linearly predicted each cognitive domain; variable selection was performed using 5-fold cross validation based on the sum of squared error criterion. Then associations of selected markers with cognitive function were assessed using linear regression models after adjusting for predictors of cognition (i.e., age, sex, education, race-ethnicity, education, insurance status, diabetes, smoking status, systolic blood pressure, diastolic blood pressure, high-density lipoprotein, low-density lipoprotein, and estimated glomerular filtration rate). The independent association of the immune marker ensemble was tested using likelihood ratio test (LRT) comparing two nested models: a model that included the traditional predictors only (M1), and a model that included the LASSO-selected markers and interactions in addition to the traditional predictors (M2). R-squared (R^2^) was used to quantify the improvement of variance in the cognitive function explained by the LASSO-selected immune marker ensemble. All analyses were performed using SAS version 9.4 (SAS Institute Inc. Cary, NC) and R version 3.4.

## RESULTS

### Description of cohort

There were 1179 participants with immune marker data available (Table 1). Mean age was 70±8.9 years and 60% were women. There was a high proportion of Hispanics (68%); mean education level was 10±5 years. There was a high burden of diabetes (28%) and other cardiovascular risk factors.

**Table 1.** Characteristics of the Cohort.

|  |  | Overall<br>N=1179 |
| --- | --- | --- |
| Age at visit 1, mean (±SD) | | 70 (±8.9) |
| Men, n (%) |  | 473 (40.1%) |
| Years of Education, mean (±SD) | | 9.7 (±5.1) |
| Race-ethnicity, n (%) | Hispanic | 785 (66.6%) |
|  | Non-Hispanic Black | 198 (16.8%) |
|  | Non-Hispanic White | 171 (14.5%) |
| Medical Insurance, n (%) | Medicaid or no Insurance | 560 (47.5%) |
|  | Medicare or Private Insurance | 619 (52.5%) |
| Current Smokers, n (%) |  | 182 (15.4%) |
| Moderate or Heavy Physical Activity, n (%) |  | 119 (10.2%) |
| Diabetes, n (%) |  | 324 (27.5%) |
| Systolic blood pressure in mm Hg, mean (±SD) | | 137 (±17.6) |
| Diastolic blood pressure in mm Hg, mean (±SD) | | 78 (±9.6) |
| High-density lipoprotein, mg/dl, mean (±SD) | | 53.3 (±16.9) |
| Low-density lipoprotein, mg/dl, mean (±SD) | | 115.6 (±35.1) |
| eGFR, mean (±SD) | | 73.6 (±18.0) |

SD=standard deviation; eGFR=estimated glomerular filtration rate

### Associations of LASSO-selected immune markers with cognitive function

Inclusion of LASSO-selected immune markers collectively improved model fit over age alone (p-value for LRT< 0.0001 for global score and for all domains) and over age, demographics including education, and traditional risk factors for each cognitive outcome (p-value <0.0001 for global score, memory, language, and executive function: p = 0.0081 for processing speed; Table 2). For global cognitive function, the variance in cognition explained by the immune markers was similar to that explained by age (model 0 and model 1-1), and the addition of immune markers to a model that included age alone led to almost a doubling of the R^2^ (model 1-1 and model 1-2), implying that the effect of the immune markers was independent and comparable in size to that of the effect of age. The effect of age alone was greatest for the domain of processing speed (R^2^ =0.17) and least for language (R^2^ =0.02). The impact of adding immune markers to age was greatest for executive function, for which R^2^ increased from 0.04 for a model including only age to 0.17 when the immune markers were added.

**Table 2.** Impact of immune markers and other factors on explanatory models for cognitive test scores.

| Cognitive function | Adjusted R<sup>2</sup> values for models containing the listed variables |  |  |  |  |
| --- | --- | --- | --- | --- | --- |
|  | Model 0 | Model 1-1 | Model 2-1 | Model 1-2 | Model 2-2 |
|  | Immune markers* only | Age only | Age, demographics and risk factors** | Immune markers* and age | Immune markers*, age, demographics, and risk factors** |
| Global Score | 0.13 | 0.12 | 0.48 | 0.21<sup>†</sup> | 0.50<sup>¥</sup> |
| Cognitive Domains |  |  |  |  |  |
| Memory | 0.12 | 0.11 | 0.31 | 0.18<sup>†</sup> | 0.33<sup>¥</sup> |
| Language | 0.09 | 0.02 | 0.39 | 0.10<sup>†</sup> | 0.41<sup>¥</sup> |
| Processing Speed | 0.11 | 0.17 | 0.31 | 0.22<sup>†</sup> | 0.32<sup>¥¥</sup> |
| Executive Function | 0.14 | 0.04 | 0.39 | 0.17<sup>†</sup> | 0.42<sup>¥</sup> |

\* Immune markers refers to those immune markers and their interactions that were selected from the Least Absolute Shrinkage and Selection Operator (LASSO) procedure as described in Methods.
\*\* Demographics and risk factors: Sex, race-ethnicity, education, medical insurance type, diabetes, smoking status, systolic blood pressure, diastolic blood pressure, high-density lipoprotein, low-density lipoprotein, estimated glomerular filtration rate
<sup>†</sup> p<0.0001 for likelihood ratio test for effect of addition of immune markers selected by LASSO to model adjusted only for age (comparing Model 1-1 and Model 1-2)
<sup>¥</sup> p<0.0001 for likelihood ratio test for effect of addition of immune markers selected by LASSO to model adjusted for age, demographics and risk factors (comparing Model 2-1 and Model 2-2)
<sup>¥¥</sup> p=0.0081 for likelihood ratio test for effect of addition of immune markers selected by LASSO to model adjusted for age, demographics and risk factors (comparing Model 2-1 and Model 2-2)

Approximately 48% of the variance in global cognition was explained by a model containing age, other demographics, and risk factors; the model fit improved by approximately 4.0% (from 48% to 50%) after inclusion of the immune markers.

### Specific immune markers predicting cognitive functions

A subset of immune markers and their two-way interactions that predicted each cognitive function were selected using LASSO (Table e-2). C-C Motif Chemokine Ligand 11 (CCL11, or eotaxin), C-X-C Motif Chemokine Ligand 9 (CXCL9), hepatocyte growth factor (HGF) and serpin E1 (or plasminogen activator inhibitor-1) were consistently selected as being associated with overall cognitive function (Table 3) and with each of the neuropsychological domains (table e-3-6). In addition, sFasL, SCF, leptin, CCL3, CSF3 and CCL2 contributed to multiple cognitive domains. Interactions between CCL5 and serpin E1, between CCL3 and TNFβ, between CCL11 and IL1RA, between HGF and leptin, between TRAIL and IL12p40, between CCL11 and CCL5, and between CXCL10 and CXCL9 were also found in 2 or more domains.

**Table 3:** Association of immune markers and interactions with global cognitive score*.

| Immune marker<sup>†</sup> | Estimate | Standard Error | p |
| --- | --- | --- | --- |
| CCL11 | -0.036 | 0.017 | 0.033 |
| HGF | -0.066 | 0.020 | 0.001 |
| CXCL9 | -0.029 | 0.017 | 0.092 |
| SCF | -0.008 | 0.018 | 0.669 |
| Leptin | 0.027 | 0.019 | 0.160 |
| sFasL | 0.043 | 0.018 | 0.015 |
| CCL3 | 0.024 | 0.019 | 0.193 |
| Serpin_E1 | 0.037 | 0.016 | 0.022 |
| Serpin_E1*CCL5 | -0.021 | 0.014 | 0.123 |
| HGF*Leptin | -0.036 | 0.017 | 0.031 |
| CCL11*IL1RA | -0.008 | 0.012 | 0.485 |
| CCL11*SCF | -0.033 | 0.015 | 0.025 |
| sFasL*PDGFBB | -0.030 | 0.015 | 0.042 |
| CCL5*CCL4 | -0.001 | 0.020 | 0.962 |
| CCL4*VCAM1 | -0.029 | 0.022 | 0.186 |
| CXCL10*IFNb | 0.009 | 0.031 | 0.761 |
| HGF*TRAIL | 0.014 | 0.017 | 0.411 |
| CXCL10*TGFa | 0.011 | 0.035 | 0.745 |
| CCL3*TNFb | 0.007 | 0.014 | 0.599 |
| TRAIL*IL12p40 | 0.025 | 0.019 | 0.179 |
| CXCL10*IL5 | -0.010 | 0.021 | 0.643 |
| CCL3*LIF | -0.006 | 0.003 | 0.670 |
| CCL11*sICAM1 | 0.020 | 0.017 | 0.243 |
| CXCL10*VEGFD | 0.013 | 0.016 | 0.432 |

\* Adjusted for age, sex, race-ethnicity, education, medical insurance type, diabetes, smoking status, systolic blood pressure, diastolic blood pressure, high-density lipoprotein, low-density lipoprotein, estimated glomerular filtration rate<sup>†</sup> Immune markers were log 2 transformed and standardized
CCL11 = C-C Motif Chemokine Ligand 11; CXCL9 = C-X-C Motif Chemokine Ligand 9; HGF = hepatocyte growth factor; sFasL = soluble Fas ligand; SCF = stem cell factor; CSF2 = Colony stimulating factor 2; CCL2 = C-C Motif Chemokine Ligand 2; CCL3 = C-C Motif Chemokine Ligand 3; CCL4 = C-C Motif Chemokine 4; IL12p40 = interleukin-12 p40; IL21 = interleukin 21; IL1a = interleukin 1a; CCL4 = C-C Motif Chemokine Ligand 4; IFNb = interferon beta; PDGFBB = Platelet derived growth factor-BB; TGFa = transforming growth factor $\alpha$ ; TRAIL = Tumor necrosis factor-related apoptosis-inducing ligand

### Magnitude of immune marker effects

The magnitude of immune marker effects was comparable to that of conventional risk factors. For example, for executive function, each SD increase in CCL11 levels (β=-0.052, p=0.016) was associated with an effect size equivalent to aging 3 years (3-year β=-0.048, p<0.0001). HGF had twice the effect on memory function (β per SD=-0.06, p=0.012) as aging (β per year =-0.03, p<0.0001).

## Discussion

We found several immune molecules are cross-sectionally associated with cognitive function in a predominantly Hispanic, but ethnically diverse, population. Many immune markers were correlated, prompting us to use the LASSO technique to perform data reduction, thereby extracting those molecules that were independently and most informatively associated with cognition. As a group, those markers that were selected using LASSO had an explanatory impact comparable and independent to that of age. After accounting for sociodemographic and vascular risk factors, as well, the immune panel significantly improved model fit for the association with cognition by approximately 4%.

These results suggest that serum immune markers have an independent association with cognition beyond that due to traditional risk factors for cognition and vascular disease. While the collective magnitude of these markers on cognition was modest, the magnitude of association for several markers was comparable to that of other well-accepted risk markers of cognitive impairment, including age. Cognitive decline is a complex, multifactorial phenomenon, and it is unlikely that any single risk factor, immune pathway or marker accounts for a substantial proportion of the variance in cognition. The total variance explained by models not including the immune panel did not exceed 50%. Our results suggest that several immune pathways may be involved and that approaches that affect multiple pathways simultaneously will be most likely to have an impact, although further research is needed.

Individual markers most consistently associated with cognition were the chemokines CCL11 and CXCL9, the neurotrophic factor HGF, and serpin E1. Chemokines are a large family of small protein molecules that play an important role in immune functions. CCL11 (eotaxin-1) primarily binds to the CCR3 receptor and is important in eosinophil recruitment, allergic responses, and skewing toward a T helper (Th)-2 immune response. Its levels increase with age, it can cross the blood brain barrier, and it has been postulated to promote aging-accelerated neurodegeneration.^23,24,25^ CCL11 impairs post-stroke recovery in animal models, although the effect in humans appears to be reversed.^26^

CXCL9 is a member of the subfamily of CXC chemokines that lack a glutamic acid-leucine-arginine motif. They all share the CXCR3 receptor, and participate in Th1-type immune responses, involving tissue infiltration by T cells as part of the innate immune response. CXCL9 is constitutively expressed on human brain-derived microvascular endothelial cells and astrocytes.^27^ CXCL9 is upregulated by IFN-γ, induces activation of extracellular signal-regulated kinases in cortical neurons in mice, and is involved in neuronal–glial interactions.^28^ The CXC chemokines increase with inflammation and infection of the CNS,^29^ and are also up-regulated in human brains with Alzheimer pathology.^30^

HGF is a neurotrophic factor with anti-apoptotic and angiogenesis effects that facilitates dendritic arborization and has been associated with small artery disease among patients with cognitive impairment and AD.^31^ Consistent with our findings, these associations were independent of other vascular risk factors, suggesting that the mechanisms by which HGF contributes to cognitive impairment may be independent of traditional vascular risk factors. It is increasingly evident that vascular and neurodegenerative contributions to cognitive aging and dementia overlap, and that vascular mechanisms contribute to neurodegeneration. Immune mechanisms that lead to small vessel disease may thus enhance the degenerative pathways that lead to AD, either by impairing blood flow, enhancing inflammation, reducing regenerative capacity, or inhibiting amyloid clearance.

Serpin E1 is the major inhibitor of the serine proteases tissue plasminogen activator and urokinase, and thus acts as an inhibitor of fibrinolysis. Elevated levels potentially contribute to increased atherosclerosis, and risk of coronary artery events and stroke.^32^ In addition, serpin E1 inhibits activity of matrix metalloproteinases, which play a role in blood-brain barrier breakdown after ischemia.^33^ Serpin E1 also plays a role in regulation of brain-derived neurotrophic factor (BDNF), which must be converted from a pro-protein by plasmin. BDNF plays an important role in neuronal activity and memory.^34^ A small study showed that serpin E1 is elevated in peripheral blood with mild cognitive impairment;^35^ however, our findings of higher levels of serpin E1 in association with cognitive performance, and without interaction with BDNF, suggest that serpin E1 production may exert a small protective effect.

Associations of immune markers with cognition were present for each of the individual domains of cognitive function investigated, although several markers, as well as a global cognitive measure, were present across all areas. Other markers selected by our data reduction technique as contributing to two or more individual cognitive domains included sFasL (memory, language function, processing speed, and global cognition), SCF (memory, processing speed, and global cognition), CSF2 (memory and processing speed), leptin (language, executive function, and global cognition), and the chemokines CCL2 (processing speed, executive function) and CCL3 (language, executive function, and global cognition).

Evidence that levels of the chemokines CCL2, CCL3, CCL11 and CXCL9 are associated with cognitive decline in our cohort support the hypothesis that the innate immune system contributes to cognitive impairment and dementia.^36,37^ Recent studies provide evidence that increased amyloid-beta peptide activates the innate immune system via pattern recognition receptors on microglia and astrocytes, contributing to hyperphosphorylation of *tau*. Sterile inflammation can also be provoked by recognition of intracellular contents released from necrotic cells through damage-associated molecular patterns. Elevations of these chemokines could reflect chronic activation of innate immunity in brains of patients with cognitive impairment. There is also evidence that infections, such as herpes simplex virus-1, provoke innate immunity, cause *tau* phosphorylation, and contribute to neurodegeneration.^38^ Previous studies in our cohort demonstrate that serologies against herpesviruses are associated with lower cognitive test scores.^39,40^

Our results could have clinical implications since inflammation is potentially modifiable. IL1β inhibition diminishes residual inflammatory risk for cardiovascular events,^41,42^ and some antiviral agents are being tested for efficacy against AD, with variable results.^43,44,45^ Acetylcholinesterase inhibitors also decreased IFNγ, TNFα, IL1β and IL6 secretion by immune cells and enhanced innate resistance to viral infection *in vitro*.^46^ Furthermore, the timing of intervention also appears to be critical, underscoring the significance of long-term prospective investigations such as NOMAS.

Our study has limitations. First, measures of immune molecules and cognition were obtained at a single time point, limiting our ability to make causal determinations. We continue to follow our cohort over time, however, to provide longitudinal data. Second, immune markers were not assessed in the cerebrospinal fluid or brain tissue, the site where injury occurs. Nonetheless, blood biomarkers are increasingly recognized as indicators of neurodegenerative activity and predictors of AD. Third, though our panel included 60 different immune-related markers, many others were not included. Fourth, we did not include MRI-derived biomarkers in this study, nor did we have positron emission tomography to assess amyloid, *tau*, or microglial activations. Our outcomes, however, were cognitive test scores, which may reflect multiple pathologies and may ultimately have greater clinical relevance for patients. Our assay does not provide a clinically useful panel of markers for the prediction of dementia currently. Our findings require confirmation in other cohorts, and in longitudinal analyses, before they can be used for clinical purposes.

Our study also has strengths. Our cohort represents a racially and ethnically diverse, elderly cohort at highest risk for cognitive decline and dementia. Many prior studies have not included large numbers of Black and Hispanic/Latino people. Our population was also well-characterized for demographic and medical risk factors. Our multiplex immune assay included a large number of molecules that may contribute to cognitive decline and dementia.

In conclusion, peripheral blood levels of several immune-related molecules, representing pathways related to innate immunity, neural repair, and thrombosis, may play a role in cognitive decline and dementia. As a group these factors modestly contribute to cognitive impairment, independently of vascular risk factors associated with cognitive decline. These results speak to the ability of a multiplex immune assay to add information regarding pathways that may be involved in neurodegeneration in a diverse population. These findings also indicate the need for further research on immune mechanisms in cognitive aging.

## Data Availability

Data are available upon appropriate request to the principal investigators.

## Non-standard Abbreviations and Acronyms

95%CI: 95% confidence interval
AD: Alzheimer Disease
CCL2: C-C Motif Chemokine Ligand 2
CCL3: C-C Motif Chemokine Ligand 3
CCL5: C-C Motif Chemokine Ligand 5
CCL11: C-C Motif Chemokine Ligand 11
CSF3: Colony stimulating factor 3
CRP: C-reactive protein
CXCL9: C-X-C Motif Chemokine Ligand 9
CXCL10: C-X-C Motif Chemokine Ligand 10
HGF: hepatocyte growth factor
IL12p40: interleukin-12 p40
IL1RA: interleukin-1 receptor antagonist
LASSO: Least Absolute Shrinkage and Selection Operator
LRT: likelihood ratio test
MCI: mild cognitive impairment
NOMAS: Northern Manhattan Study
SCF: stem cell factor
sFasL: soluble Fas ligand
TNFβ: tumor necrosis factor β
TRAIL: tumor necrosis factor-related apoptosis-inducing ligand

