## Supplemental Tables for "Immune Markers Are Associated with Cognitive Performance in a Multiethnic Cohort: the Northern Manhattan Study"

### Table e-1

| Cognitive domain | Assessments |
|---|---|
| Memory | Three sub-scores derived from a 12-word five trial list-learning task: list learning total score, delayed recall score, and delayed recognition score |
| Language | Picture naming (modified Boston Naming) test, Category fluency (Animal Naming) test, Phonemic fluency (C, F, L in English speakers and P, S and V in Spanish speakers) |
| Processing Speed | Color Trails test Form 1[¥],<br>Grooved Pegboard task (non-dominant hand) [¥] |
| Executive Function | Difference in time to complete the Color Trails test Form 1[¥] and Form 2[¥],<br>Sum of the Odd-Man-Out subtests 2 and 4 |

[¥] For the participants who did not complete the tasks within the maximum time allowed, “imputed” time was calculated by adding penalty time to maximum time allowed. The penalty time was obtained by multiplying the number of incomplete items by “expected time per correct items completed” estimated using Kaplan-Meier.

**Table e-2: immune markers selected by Least Absolute Shrinkage and Selection Operator procedure as linearly associated with global cognition score and each domain score**

| Global Cognitive Score | Memory | Language | Processing Speed | Executive Function |
|---|---|---|---|---|
| CCL11 | CCL11 | CCL11 | CCL11 | CCL11 |
| CXCL9 | CXCL9 | CXCL9 | CXCL9 | CXCL9 |
| HGF | HGF | HGF | HGF | HGF |
| Serpin_E1 | Serpin_E1 | Serpin_E1 | Serpin_E1 | Serpin_E1 |
| sFasl | sFasl | sFasl | sFasl | Leptin |
| SCF | CSF2 | Leptin | CSF2 | CCL3 |
| Leptin | SCF | CCL3 | SCF | CCL2 |
| CCL3 | CXCL12a | IL12p40 | CCL2 | PDGFBB |
|  | IL21 |  | IL1a |  |
|  | CCL4 |  | IFNb |  |
|  | TGFa |  |  |  |

CCL11 = C-C Motif Chemokine Ligand 11; CXCL9 = C-X-C Motif Chemokine Ligand 9; HGF = hepatocyte growth factor; sFasL = soluble Fas ligand; SCF = stem cell factor; CSF2 = Colony stimulating factor 2; CCL2 = C-C Motif Chemokine Ligand 2; CCL3 = C-C Motif Chemokine Ligand 3; CCL4 = C-C Motif Chemokine 4; IL12p40 = interleukin-12 p40; IL21 = interleukin 21; IL1a = interleukin 1a; CCL4 = C-C Motif Chemokine Ligand 4; IFNb = interferon beta; PDGFBB = Platelet derived growth factor-BB; TGFa = transforming growth factor α; TRAIL = Tumor necrosis factor-related apoptosis-inducing ligand

**Table e-3. Association of immune markers and interactions with memory domain***

| Immune marker‡ | Estimate | Standard Error | Pr > ChiSq |
|---|---|---|---|
| CXCL9 | -0.047 | 0.03 | 0.115 |
| HGF | -0.082 | 0.029 | 0.005 |
| CCL11 | 0.005 | 0.025 | 0.844 |
| SCF | -0.050 | 0.027 | 0.061 |
| CSF2 | 0.057 | 0.029 | 0.051 |
| Serpin_E1 | 0.057 | 0.025 | 0.021 |
| IL21 | -0.011 | 0.074 | 0.880 |
| sFasL | 0.029 | 0.026 | 0.262 |
| CCL4 | 0.126 | 0.066 | 0.055 |
| TGFa | 0.019 | 0.028 | 0.504 |
| TGFa*CXCL10 | -0.011 | 0.045 | 0.814 |
| CCL11*CCL5 | 0.031 | 0.023 | 0.179 |
| CCL2*IL12p40 | 0.007 | 0.018 | 0.693 |
| CXCL10*TNFa | 0.015 | 0.025 | 0.547 |
| sFasL*PDGFBB | -0.042 | 0.021 | 0.043 |
| TRAIL*VCAM1 | -0.062 | 0.026 | 0.015 |
| CXCL9*CXCL10 | -0.026 | 0.018 | 0.144 |
| HGF*Leptin | -0.035 | 0.024 | 0.145 |
| CXCL12a | -0.041 | 0.070 | 0.556 |
| CXCL10*IFNb | 0.033 | 0.044 | 0.456 |
| IL12p40*TRAIL | 0.039 | 0.022 | 0.071 |

*Adjusted for age, sex, race-ethnicity, education, medical insurance type, diabetes, smoking status, systolic blood pressure, diastolic blood pressure, high-density lipoprotein, low-density lipoprotein, estimated glomerular filtration rate
‡ Immune markers were log 2 transformed and standardized

CCL11 = C-C Motif Chemokine Ligand 11; CXCL9 = C-X-C Motif Chemokine Ligand 9; HGF = hepatocyte growth factor; sFasL = soluble Fas ligand; SCF = stem cell factor; CSF2 = Colony stimulating factor 2; CCL2 = C-C Motif Chemokine Ligand 2; CCL3 = C-C Motif Chemokine Ligand 3; CCL4 = C-C Motif Chemokine 4; IL12p40 = interleukin-12 p40; IL21 = interleukin 21; IL1a = interleukin 1a; CCL4 = C-C Motif Chemokine Ligand 4; IFNb = interferon beta; PDGFBB = Platelet derived growth factor-BB; TGFa = transforming growth factor α; TRAIL = Tumor necrosis factor-related apoptosis-inducing ligand

**Supplement Table e-4. Association of immune markers and interactions with language domain***

| Immune marker[ǂ] | Estimate | Standard Error | p |
|---|---|---|---|
| IL12p40 | -0.066 | 0.029 | 0.024 |
| HGF | -0.045 | 0.029 | 0.126 |
| CCL11 | -0.038 | 0.021 | 0.073 |
| Serpin_E1 | 0.012 | 0.026 | 0.641 |
| sFasL | 0.055 | 0.022 | 0.012 |
| VCAM1 | 0.044 | 0.027 | 0.100 |
| Leptin | 0.046 | 0.024 | 0.058 |
| CXCL9 | -0.015 | 0.024 | 0.546 |
| CCL3 | 0.020 | 0.023 | 0.398 |
| IL7*PDGFBB | -0.016 | 0.024 | 0.490 |
| CXCL9*CXCL10 | -0.009 | 0.016 | 0.566 |
| PDGFBB*IL13 | -0.013 | 0.025 | 0.598 |
| Serpin_E1*CCL5 | -0.022 | 0.018 | 0.233 |
| IL12p40*TRAIL | 0.027 | 0.017 | 0.114 |
| CCL3*TNFb | -0.003 | 0.012 | 0.834 |
| CXCL10*CCL5 | 0.033 | 0.020 | 0.102 |

*Adjusted for age, sex, race-ethnicity, education, medical insurance type, diabetes, smoking status, systolic blood pressure, diastolic blood pressure, high-density lipoprotein, low-density lipoprotein, estimated glomerular filtration rate
ǂ Immune markers were log 2 transformed and standardized

CCL11 = C-C Motif Chemokine Ligand 11; CXCL9 = C-X-C Motif Chemokine Ligand 9; HGF = hepatocyte growth factor; sFasL = soluble Fas ligand; SCF = stem cell factor; CSF2 = Colony stimulating factor 2; CCL2 = C-C Motif Chemokine Ligand 2; CCL3 = C-C Motif Chemokine Ligand 3; CCL4 = C-C Motif Chemokine 4; IL12p40 = interleukin-12 p40; IL21 = interleukin 21; IL1a = interleukin 1a; CCL4 = C-C Motif Chemokine Ligand 4; IFNb = interferon beta; PDGFBB = Platelet derived growth factor-BB; TGFa = transforming growth factor α; TRAIL = Tumor necrosis factor-related apoptosis-inducing ligand

**Supplement Table e-5. Association of immune markers and interactions with processing speed***

| Immune marker‡ | Estimate | Standard Error | p |
|---|---|---|---|
| CXCL9 | -0.034 | 0.032 | 0.297 |
| CCL11 | -0.045 | 0.026 | 0.086 |
| HGF | -0.071 | 0.031 | 0.023 |
| CSF2 | 0.010 | 0.031 | 0.739 |
| CCL2 | 0.000 | 0.026 | 0.999 |
| SCF | -0.007 | 0.029 | 0.804 |
| IL1a | 0.011 | 0.033 | 0.740 |
| IFNb | 0.046 | 0.029 | 0.119 |
| sFasL | 0.019 | 0.028 | 0.492 |
| Serpin_E1 | 0.033 | 0.024 | 0.181 |
| HGF*Leptin | -0.060 | 0.027 | 0.028 |
| CCL11*IL1RA | -0.049 | 0.017 | 0.005 |
| Serpin_E1*CCL5 | -0.022 | 0.022 | 0.313 |
| Leptin*VEGFD | -0.047 | 0.026 | 0.066 |
| Leptin*CCL4 | -0.032 | 0.031 | 0.289 |
| CXCL9*IL10 | -0.030 | 0.023 | 0.190 |
| CSF2*IL17F | 0.092 | 0.063 | 0.142 |
| VEGFD*CXCL10 | 0.056 | 0.024 | 0.021 |
| CCL11*sICAM1 | 0.039 | 0.028 | 0.157 |
| Leptin*CXCL10 | 0.053 | 0.050 | 0.294 |
| HGF*CXCL12a | 0.028 | 0.020 | 0.159 |
| SCF*BDNF | -0.002 | 0.026 | 0.931 |
| CCL2*BDNF | 0.022 | 0.023 | 0.352 |
| IFNb*PDGFBB | 0.017 | 0.025 | 0.500 |
| IL10*BDNF | 0.004 | 0.036 | 0.908 |
| Serpin_E1*IL31 | 0.054 | 0.025 | 0.028 |
| IL10*PDGFBB | 0.026 | 0.031 | 0.404 |
| IL1RA*BDNF | 0.012 | 0.029 | 0.669 |
| CCL11*CCL5 | 0.023 | 0.026 | 0.372 |
| CCL3*TNFb | -0.012 | 0.016 | 0.458 |
| IFNb*CXCL10 | -0.011 | 0.028 | 0.685 |
| CSF2*CXCL10 | -0.027 | 0.054 | 0.620 |
| CSF2*IFNb | -0.045 | 0.058 | 0.438 |

*Adjusted for age, sex, race-ethnicity, education, medical insurance type, diabetes, smoking status, systolic blood pressure, diastolic blood pressure, high-density lipoprotein, low-density lipoprotein, estimated glomerular filtration rate‡ Immune markers were log 2 transformed and standardized

CCL11 = C-C Motif Chemokine Ligand 11; CXCL9 = C-X-C Motif Chemokine Ligand 9;
HGF = hepatocyte growth factor; sFasL = soluble Fas ligand; SCF = stem cell factor;
CSF2 = Colony stimulating factor 2; CCL2 = C-C Motif Chemokine Ligand 2;
CCL3 = C-C Motif Chemokine Ligand 3; CCL4 = C-C Motif Chemokine 4;

IL12p40 = interleukin-12 p40; IL21 = interleukin 21; IL1a = interleukin 1a; CCL4 = C-C Motif Chemokine Ligand 4; IFNb = interferon beta; PDGFBB = Platelet derived growth factor-BB; TGFa = transforming growth factor α; TRAIL = Tumor necrosis factor-related apoptosis-inducing ligand

**Supplement Table e-6. Association of immune markers with executive function domain***

| Immune marker‡ | Estimate | Standard Error | p |
|---|---|---|---|
| CCL11 | -0.056 | 0.023 | 0.014 |
| Leptin | -0.005 | 0.026 | 0.859 |
| HGF | -0.075 | 0.026 | 0.004 |
| CXCL9 | 0.026 | 0.026 | 0.319 |
| CCL2 | -0.011 | 0.022 | 0.619 |
| CCL3 | 0.050 | 0.025 | 0.047 |
| Serpin_E1 | 0.030 | 0.022 | 0.172 |
| PDGFBB | 0.058 | 0.021 | 0.006 |
| CCL11*SCF | -0.039 | 0.021 | 0.057 |
| Serpin_E1*CCL5 | -0.038 | 0.018 | 0.037 |
| CCL4*VCAM1 | -0.050 | 0.030 | 0.099 |
| CCL5*CCL4 | 0.012 | 0.028 | 0.656 |
| Leptin*CXCL12a | -0.016 | 0.028 | 0.562 |
| Leptin*IFNa2 | -0.027 | 0.023 | 0.248 |
| sFasL*IL21 | 0.001 | 0.015 | 0.924 |
| IL21*IL7 | 0.017 | 0.019 | 0.376 |
| CCL4*VEGFA | -0.016 | 0.010 | 0.098 |
| VEGFA*IL13 | -0.040 | 0.017 | 0.023 |
| CCL11*IL1RA | -0.013 | 0.016 | 0.425 |
| PDGFBB*BDNF | -0.023 | 0.020 | 0.250 |
| IL5*CXCL10 | -0.065 | 0.087 | 0.458 |
| PDGFBB*VEGFD | 0.043 | 0.022 | 0.048 |
| CCL3*TNFb | -0.001 | 0.018 | 0.950 |
| CXCL9*PDGFBB | 0.040 | 0.029 | 0.055 |
| CCL3*sICAM1 | 0.052 | 0.025 | 0.039 |
| Serpin_E1*IL17F | 0.025 | 0.020 | 0.215 |
| sICAM1*TNFa | 0.008 | 0.024 | 0.730 |
| CXCL10*IL22 | -0.001 | 0.044 | 0.974 |
| CXCL10*IL2 | 0.078 | 0.081 | 0.336 |
| CCL3*IL5 | 0.042 | 0.022 | 0.056 |

*Adjusted for age, sex, race-ethnicity, education, medical insurance type, diabetes, smoking status, systolic blood pressure, diastolic blood pressure, high-density lipoprotein, low-density lipoprotein, estimated glomerular filtration rate
‡ Immune markers were log 2 transformed and standardized

CCL11 = C-C Motif Chemokine Ligand 11; CXCL9 = C-X-C Motif Chemokine Ligand 9;
HGF = hepatocyte growth factor; sFasL = soluble Fas ligand; SCF = stem cell factor;
CSF2 = Colony stimulating factor 2; CCL2 = C-C Motif Chemokine Ligand 2;
CCL3 = C-C Motif Chemokine Ligand 3; CCL4 = C-C Motif Chemokine 4;
IL12p40 = interleukin-12 p40; IL21 = interleukin 21; IL1a = interleukin 1a; CCL4 = C-C Motif Chemokine Ligand 4; IFNb = interferon beta; PDGFBB = Platelet derived growth factor-BB;

TGFa = transforming growth factor α; TRAIL = Tumor necrosis factor-related apoptosis-inducing ligand
